# IMMUNE PROFILING UNCOVERS POTENT ADJUVANT CAPACITIES OF SARS-COV-2 INFECTION TO VACCINATION LEADING TO MEMORY T CELL RESPONSES WITH A TH17 SIGNATURE IN CANCER PATIENTS

**DOI:** 10.1101/2022.05.27.22275672

**Authors:** Miriam Echaide, Ibone Labiano, Marina Delgado, Angela Fernández de Lascoiti, Patricia Ochoa, Maider Garnica, Pablo Ramos, Luisa Chocarro, Leticia Fernández, Hugo Arasanz, Ana Bocanegra, Ester Blanco, Sergio Piñeiro, Ruth Vera, Maria Alsina, David Escors, Grazyna Kochan

**Author notes:** Corresponding authors: David Escors, Grazyna Kochan, Navarrabiomed, Irunlarrea 3, 31008 Pamplona, Navarra, Spain.

## Abstract

It is unclear whether cancer patients show impaired responses to COVID-19 and vaccination. Immune profiling was performed in three cohorts of healthy donors and oncologic patients: infected with SARS CoV-2, BNT162b2-vaccinated, and with previous COVID-19 and subsequently vaccinated. Vaccination was a poor inductor of T cell responses compared to infection, which significantly potentiated vaccination in antibody and T cell responses. T cell major targets in natural infection were the M and S protein, but not the N protein. T cell responses quickly decayed after 6 months post-vaccination, and T cell profiling showed that vaccination expanded effector T cells rather than memory T cell subsets unless the subjects had previous COVID-19. Cancer patients with previous COVID-19 and vaccinated exhibited potent IL-17+ CD4 and CD8 responses and increased neutrophils. Concluding, COVID-19 infection had potent adjuvant effects for vaccination leading to memory T cell differentiation, but with enhanced IL-17 inflammation signatures.

**Teaser:** Adjuvancy of SARS CoV-2 in cancer patients.

## Introduction

Severe acute respiratory syndrome coronavirus-2 (SARS CoV-2) caused a new outbreak of pneumonia in Wuhan, China, in December 2019(1). Since then, it has caused the COVID-19 pandemic (2). Patients with cancer are thought to be at higher risk of contracting a severe disease leading to death (3). Patients with cancer often present co-morbidities and risk factors associated with COVID-19 severity, including older age, chronic inflammation and genetic alterations associated with severe disease (2-5). Patients with cancer are usually immunocompromised by the disease and antineoplastic treatments (3, 6-8). Another frequent feature in cancer patients is T cell senescence. During aging, T cells proceed towards terminal differentiation by a sequential loss of CD27 and CD28 co-receptor surface expression (9, 10). T cell senescence is characterized loss of effector functions and impaired anti-viral immunity. Senescent T cells are enriched in effector phenotypes such as effector-memory (CD62L-CD45RA-) and effector T cells (CD62L-CD45RA+), with a loss of central memory (CD62L+ CD45RA-) and naïve (CD62L+ CD45RA+) phenotypes (10). On top of this, cancers can exacerbate chronic inflammation, which may favour pro-inflammatory cytokine release that may contribute to COVID-19 clinical syndrome (11, 12). It is yet unclear who these alterations impact immunity against SARS-CoV-2 and responses to vaccination (3, 13, 14).

Immune responses to SARS CoV-2 infection in healthy subjects are diverse and complex (15, 16), and only few studies have addressed this in cancer patients. In general, oncologic patients have shown comparable antibody responses (13, 17-19). However, T cell responses were strongly reduced in oncologic patients (17). Although regarded as a risk population, cancer patients were underrepresented in clinical trials assessing vaccine safety and efficacy (20, 21). Overall, high seroconversion rates were shown with comparable or slightly lower antibody titres in patients with solid tumours compared to healthy donors (19, 20, 22-34). However, a meta-analysis of 35 studies suggested lower protection by vaccination in oncologic patients (35). T cell activities towards the S protein (22, 23, 29, 33) showed from insufficient responses (22), lower activation rates (23, 33) or comparable to healthy donors (29).

So far, detailed information on SARS CoV-2 immunity and responses to vaccination in patients with cancer is still lacking (14, 35). For instance, three of the structural proteins (S, M and N) are the main components of the coronavirion (36, 37), but only the S protein is included in most vaccine formulations in Europe. Therefore, immune responses towards other structural proteins remain poorly studied (38, 39). Finally, it is still far from clear whether previous infection affects the responses to vaccination in cancer patients, particularly in T cell immunity.

## Results

### Cohort characteristics

Clinical and SARS-CoV-2-related characteristics of the cohorts are summarized in Table 1 and Table 2. Most oncologic (O) patients with solid tumours were under anti-neoplastic treatments, mostly chemotherapy at the time of sample collection. Treatments were not interrupted during vaccination. The degree of COVID-19 severity was generally higher in O patients compared to healthy (H) donors (fig. S2A). Disease severity was significantly different between H donors with previous COVID-19 disease (H-CoV) and O patients with previous COVID-19 disease (O-CoV) (fig. S2B). The majority of donors were vaccinated with BNT162b2 (Pfizer), inducing mild adverse events in 5% of O patients. The time elapsed from SARS CoV-2 infection or vaccination to sample collection was heterogeneous (fig. S2, D and E), but only significantly different in O-CoV versus vaccinated oncologic patients with previous COVID-19 disease (O-CoV-V), which was longer for the latter group. 45.5% of vaccinated healthy (H-V) donors completed the vaccination regime more than 6 months before sample extraction.

**Table 1.**
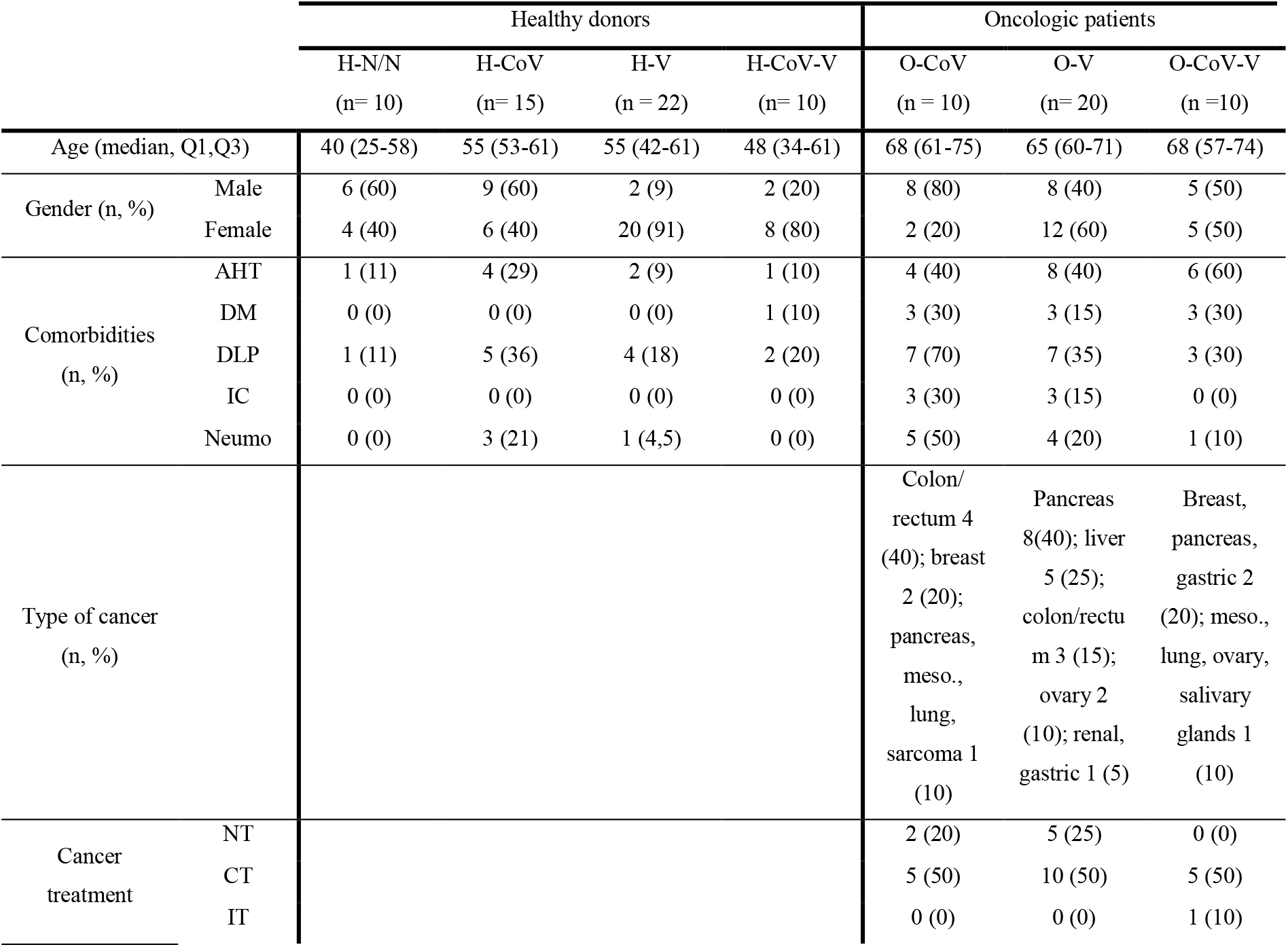
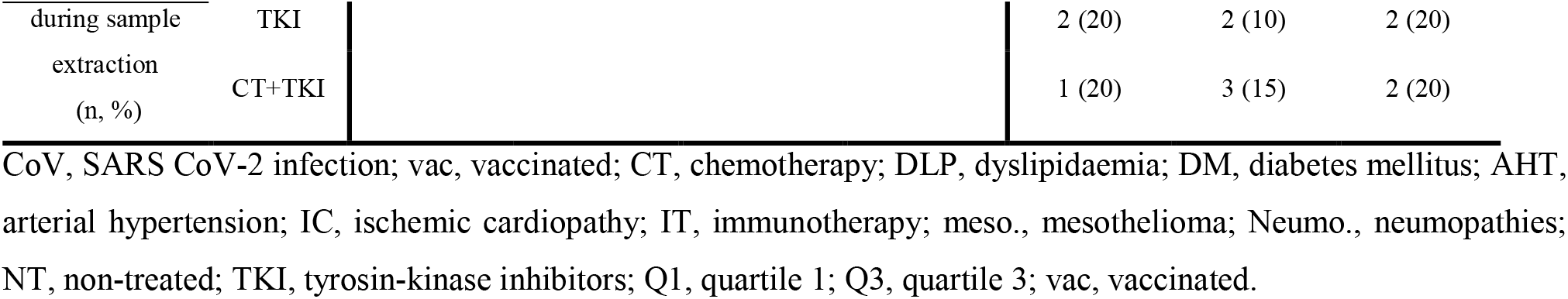
Clinical and demographic characteristics of the study cohort.

**Table 2.**
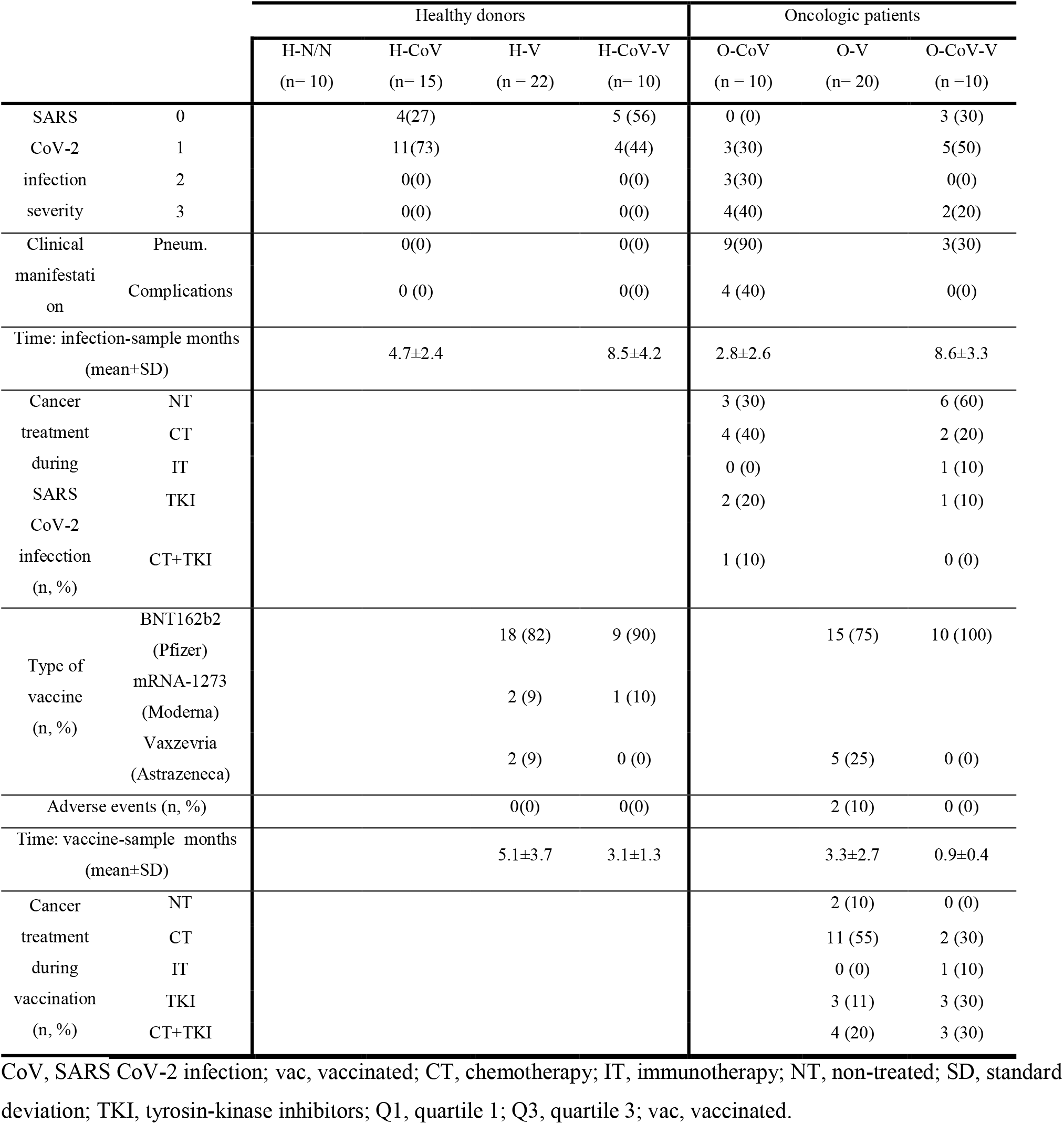
SARS CoV-2- related parameters of the study cohort.

Before all the analyses, we confirmed no significant differences in antibody titres, CD4 or CD8 T cell responses between women and men after vaccination. No significant interaction was found between sex and vaccination.

### Profiling of antibody responses towards S, M and N proteins

IgG antibody responses were evaluated towards SARS CoV-2 infection and vaccination. IgG antibody titres towards the viral proteins S, M and N were quantified, and sera from pre-pandemic donors served as technical negative controls (TC).

Infected healthy and oncologic individuals (H-CoV and O-CoV) had low S-specific IgG titres (42). No differences were observed as well between H-N/N (healthy no COVID, no vaccinated) and H-CoV groups (Fig. 1, A and B). As expected, vaccination significantly increased titres in H donors and O patients (H-V and O-V groups) (Fig. 1, A and B).

**Fig. 1.**
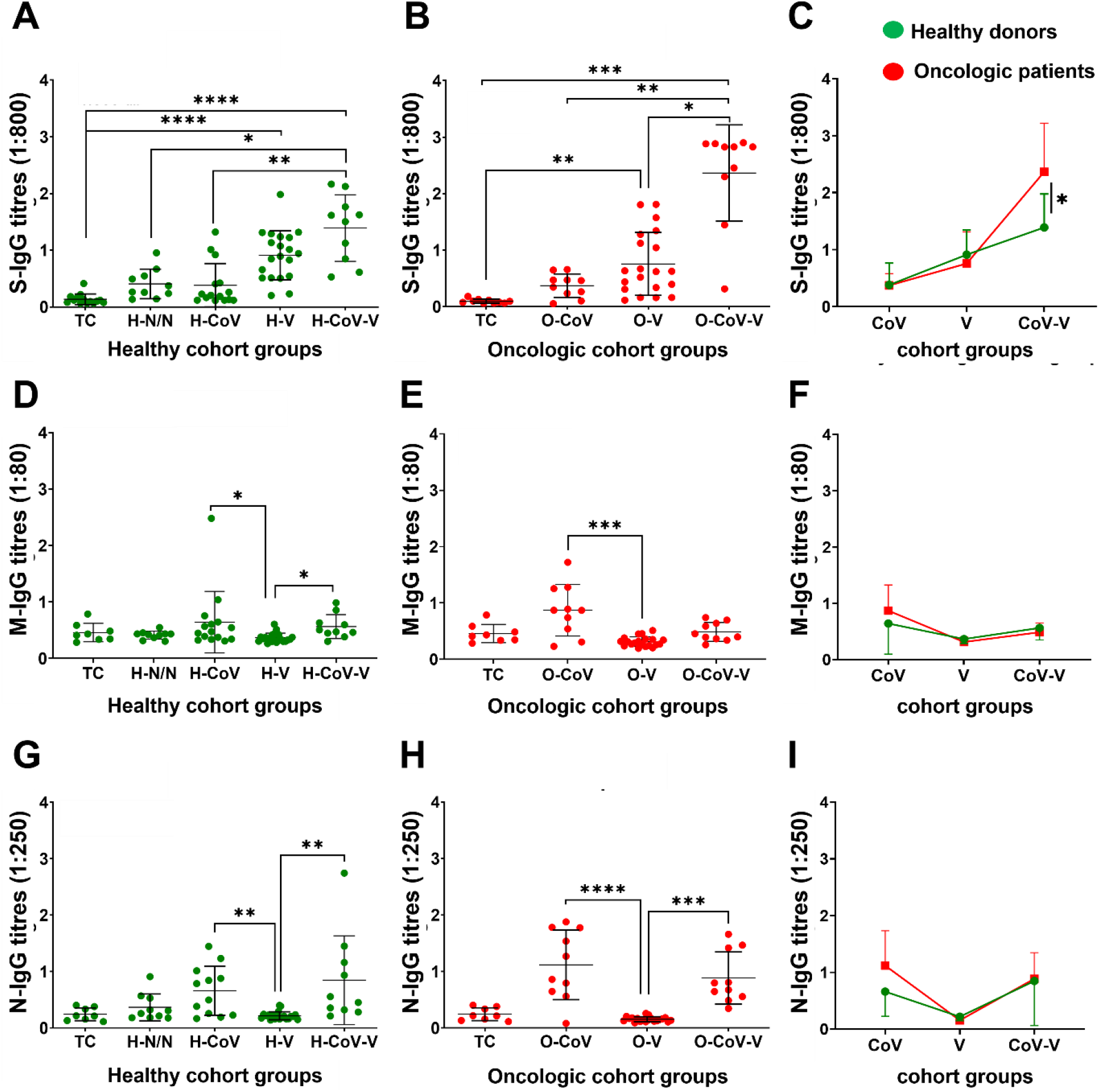
IgG antibody titres against S, M and N proteins. **(A, B, C)** S-specific IgG antibody titres in sera (1:800) from H donors and O patients. (D, E, F) M-specific IgG antibody titres in sera (1:80) from H donors and O patients. (G, H, I) N-specific IgG antibody titres in sera (1:250) from H donors and O patients. Non-parametric Krustal-Wallis test was used for multiple comparisons followed by Dunn’s test for selected pair-wise comparisons. *, **, *** and **** indicate a p value <0.05, <0.01, <0.001 and <0.0001, respectively.

Vaccination in previously infected subjects (H-CoV-V and O-CoV-V) elevated S-specific IgG titers highly significantly, suggesting a potent adjuvant effect over vaccination (Fig. 1, A and B). Interestingly, S-specific antibody titres were elevated in O-CoV-V group compared to H-CoV-V group (Fig. 1C). A positive correlation between antibody titres and disease severity was described before (16). However, the differences in COVID-19 severity between O-CoV-V and H-CoV-V groups were not significant (fig. S2C). Otherwise, the time elapsed from vaccination to sample collection was shorter in O-CoV-V compared to H-CoV-V (fig. S2E). To find out if this could be the case, IgG titres were quantified in H-V donors as a function of the time elapsed from vaccination to sample collection. There was a non-significant reduction of IgG titres in donors who completed their vaccination regimen more than 6 months before sample collection (fig. S3).

M-specific IgG antibody titres were generally very low, and previous SARS CoV-2 or vaccination did not have an effect with the exception of a mild elevation in H-CoV and O-CoV groups (Fig. 1, D and E). No differences were found between H donors and O patients (Fig. 1F). In contrast, N-specific IgG titres in infected groups (H-CoV, H-CoV-V, O-CoV and O-CoV-V) were significantly elevated compared to their vaccinated counterparts (H-V and O-H) (Fig. 1, G and H), without differences between H donors and O patients (Fig. 1I).

### Profiling of CD4 T cell activation and differentiation phenotypes

Specific T cell responses towards the three main structural proteins were evaluated in peripheral blood mononuclear cells (PBMCs). PBMCs were incubated with viral protein-specific peptivators and upregulation of the early activation markers CD154 and CD137 assessed by flow cytometry. Non-stimulated PBMCs were used as a control (NST) (fig. S4). S-specific CD4 T cells were detectable in patients with previous SARS-CoV-2 infection, especially in O patients. However, vaccination alone was not a potent inductor of S-specific CD4 T cells. In contrast, CD4 T cell responses were boosted in CoV-V groups, suggesting again a potent adjuvant effect of previous infection over T cell responses as well (Fig. 2, A and B). No differences were observed between H donors and O patients (Fig. 2C).

**Fig. 2.**
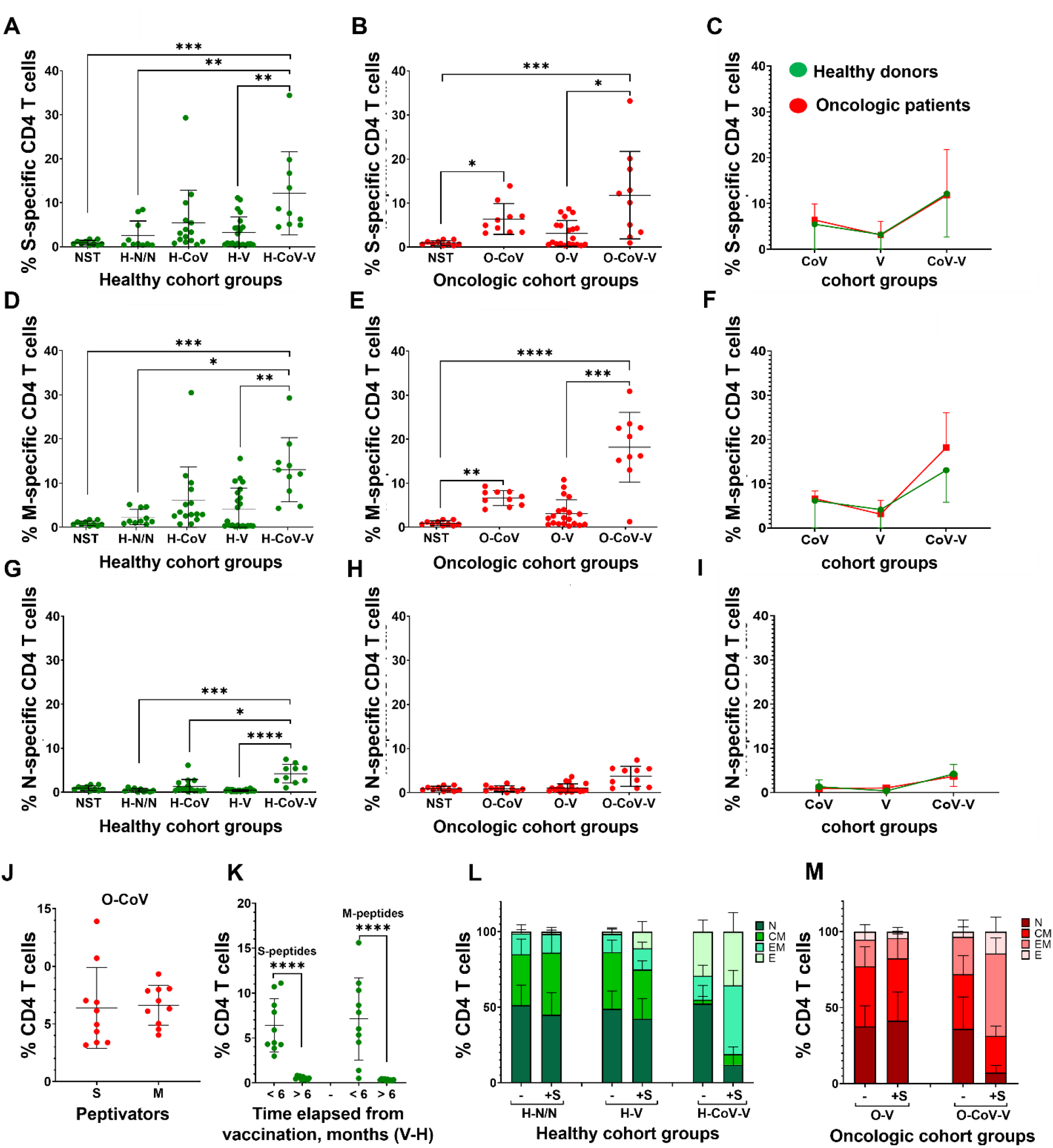
CD4 T cell responses to S, M and N peptides of SARS CoV-2 proteins. **(A, B, C)** Percentage CD4 T cells in PBMCs stimulated with S peptides in H donors and O patients. (D, E, F) Percentage CD4 T cells in PBMCs stimulated with M peptides in H donors and O patients. (G, H, I) Percentage CD4 T cells in PBMCs stimulated with N peptides in H donors and O patients. (A-I) Statistical significance was tested by Krustal-Wallis tests followed by Dunn’s pair-wise comparison tests. (J) Dot plot of the percentage of S and M-specific CD4 T cells. The U of Mann-Whitney was used to test differences. (K) Dot plot of S- and M-specific CD4 T cells in vaccinated H donors, from samples collected at the indicated timelines. Pair-wise comparisons were performed using the U of Mann-Whitney. (L,M) Relative percentages of CD4 T cell differentiation phenotypes in H donors and O patients. N, CM, EM and E, indicate naïve-stem cell (CD62L+ CD45RA+), central memory (CD62L+ CD45RA-), effector memory (CD62L-CD45RA-) and effector (CD62L-CD45RA+) phenotypes. NST, technical control of non-stimulated PMBCs. Relevant statistical comparisons are detailed in fig. S6 B-F. *, **, ***, **** indicate P <0.05, <0.01, <0.001 and <0.0001, respectively.

Similar results were found for M-specific CD4 T cells, which were the most abundant in donors with previous COVID-19 and similar in H and O patients (H-CoV-V and O-CoV-V) (Fig. 2, D-F). Unexpectedly, some CD4 reactivity towards M protein was observed in H-V and O-V donors, who did not have a previous infection. These results suggested expansion of cross-reactive CD4 T cells specific possibly towards other human coronaviruses caused by vaccination. Indeed, the M protein sequence is well-conserved between human coronaviruses (fig. S5).

CD4 responses towards the N protein were only detected in vaccinated donors with previous infection (H-CoV-V and O-CoV-V). We found that N is a poor inductor of T cell responses both in H and O donors, requiring natural infection and vaccination to expand them (Fig. 2, G-I). Both S and M proteins were equally good inductors of CD4 responses (figure 2j), but significantly decayed after 6 months post-vaccination (Fig. 2K).

Most oncologic patients have dysfunctional T cell immunity with altered T cell phenotypes (42). To investigate if this was the case after infection or vaccination, CD62L and CD45RA expression profiles were characterized in S-specific CD4 T cells (fig. S6A). Vaccination alone caused a significant increase in effector cells (CD62L-CD45RA+) in H-V donors (Fig. 2L and fig. S6B). Importantly, H and O donors with previous COVID-19 showed a significant increase in effector memory (CD62L-CD45RA-) and effector (CD62L-CD45RA+) S-specific CD4 T cells following vaccination, compared to their counterparts without COVID-19 infection. These results showed that SARS-CoV-2 infection was a potent adjuvant to vaccination in addition to expanding memory T cells (Fig. 2, L and M, and fig. S6, C-F). No significant differences were observed in CD27/CD28 expression profiles in T cells within H and O groups (fig. S6, G and H).

### Profiling of CD8 T cell activation and differentiation phenotypes

The strongest inducer of CD8 T cell responses was having a previous infection with COVID-19 both for O and H donors but not vaccination alone (Fig. 3, A-C and fig. S7). There was a tendency to increased percentages of S-specific CD8 T cells in O patients which could be caused by the differences in COVID-19 severity (fig. S2B). A similar pattern of response was observed for M-specific CD8 T cells (Fig. 3, D and E). No evidence of potential cross-reactive M-specific CD8 T cells was observed after vaccination in subjects without previous COVID-19. A tendency to elevated M-specific CD8 T cells in O-CoV group was found compared to H-CoV (Fig. 3F and fig. S2B).

**Fig. 3.**
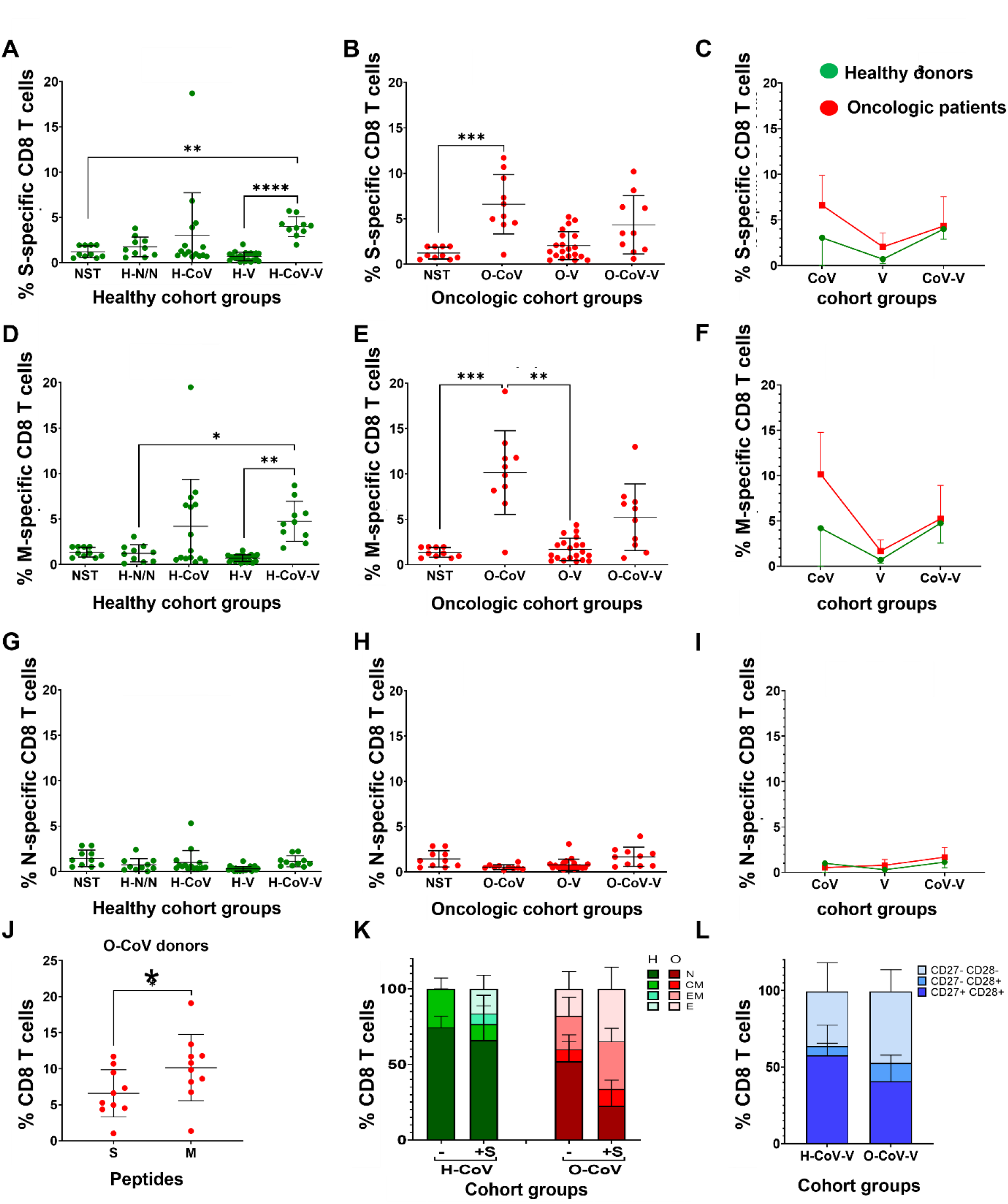
SARS-CoV-2-CD8 T cell responses in H and O donors. **(A, B, C)** Percentage of S-specific CD8 T cells in PBMCs stimulated with S-peptides in H donors and O patients as indicated. (D, E, F) Percentage of S-specific CD8 T cells in PBMCs stimulated with M-peptides in H donors and O patients. (G, H, I) Percentage of S-specific CD8 T cells in PBMCs stimulated with N-peptides in H donors and O patients. (A-I) Significance was tested with Krustal-Wallis followed by Dunn’s test. (J) Percentage of activated CD8 T cells after stimulation with S or M specific peptides in O-CoV donors. U of Mann-Whitney was used to test for significance. (K) Relative percentages of CD8 T cell differentiation phenotypes in the indicated groups of H and O donors. Means and error bars (standard deviations) are shown. N, CM, EM and E, indicate naïve-stem cell (CD62L+ CD45RA+), central memory (CD62L+ CD45RA-), effector memory (CD62L-CD45RA-) and effector (CD62L-CD45RA+) phenotypes. Relevant statistical differences are detailed in fig. S8J) Relative percentages of CD8 T cell differentiation phenotypes in the indicated groups of H and O donors. CD27+ CD28+, CD27-CD28+ and CD27+ CD28+ indicate poorly differentiated, intermediate differentiated and highly differentiated T cell phenotypes.. Relevant statistical differences are detailed in fig. S8. *, ** and *** indicate significant (P<0.05), very significant (P<0.01) and highly significant (P<0.001) differences. NST, technical control of non-stimulated (NST) PMBCs.

No N-specific CD8 responses were observed (Fig. 3, G-I). While the N protein was found a poor inductor of CD8 T cells, the M protein was instead the main target as evaluated in the O-CoV group (Fig. 3J).

No differences in S-specific CD8 phenotypes or changes were observed between H-V and O-V (fig. S8, A-F). However, there were marked baseline differences between H-CoV-V and O-CoV-V who had COVID-19 infection (Fig. 3K, fig. S8, B-G). A large proportion of S-specific CD8 T cells in H-CoV-V were poorly differentiated phenotypes (CD62L+ CD45RA+) before stimulation with S peptides. Their oncologic counterparts had expanded effector memory and effector T cell compartments (Fig. 3K and fig. S8H). After stimulation with S peptides, in contrast to H donors, O patients further expanded T cells with effector phenotypes with a drastic reduction of naïve T cells (Fig. 3K and fig. S7I). These results strongly indicated that O-CoV-V donors had exacerbated effector memory and effector responses. Indeed, baseline T cell phenotypes in O-CoV-V patients showed an drastic reduction in poorly differentiated (CD27+ CD28+) CD8 T cells compared to the non-oncologic H-CoV-V counterparts (Fig. 3L, and fig. S8J). Not significant differences were found between H-V and O-V (fig. S8K).

### Evaluation of inflammatory cytokine expression in T cells

As severe COVID-19 is associated with exacerbated inflammatory responses, IFNγ and IL-17 expression was evaluated first within S and M-specific CD4 T cells following stimulation (fig. S9). Overall, the proportion of inflammatory CD4 T cells was heterogeneous. Previous infection induced INFγ-CD4 T cells in H donors and O patients (H-CoV and O-CoV), significantly higher in O-CoV compared to O-V group. In H donors, the CoV-V group showed the strongest responses (Fig. 4, A and B). Nevertheless, responses were comparable between H donors and O patients (Fig. 4C). Similar results were observed for M-specific IFNg-CD4 T cells (Fig. 4, D-F).

**Fig. 4.**
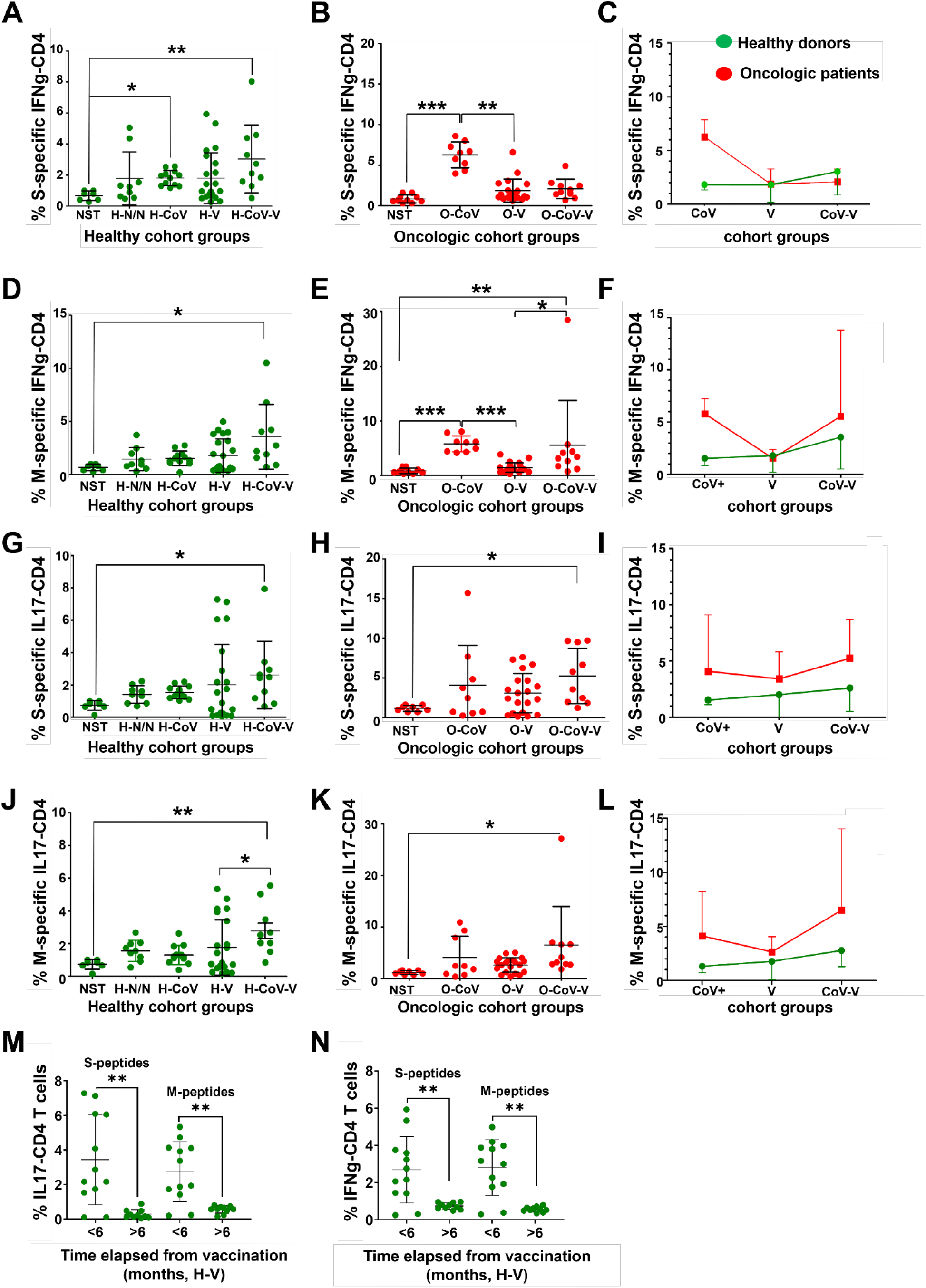
IFNγ and IL-17 expression in CD4 T cells specific for S and M proteins. **(A, B, C)** Percentage of IFNγ-CD4 T cells specific for the S protein in H and O groups. (D, E, F) Percentage of IFNγ-CD4 T cells specific for the M protein in H and O groups. (G, H, I) Percentage of IL-17-CD4 T cells specific for the S protein in H and O groups. (J, K, L) Percentage of IL-17-CD4 T cells specific for the M protein in H and O groups. (A-L) Statistical significance was evaluated with Kruskal-Wallis followed by Dunn’s pair-wise comparisons. (M, N) Percentage of CD4 T cells expressing IL-17 and IFNγ expression in H-V donors that completed the vaccine regime before sample collection in the indicated timelines, for both S and M proteins. Significance was tested with the U of Mann-Whitney test. NST, technical control of non-stimulated PMBCs. *, **, *** indicate significant (P<0.05), very significant (P<0.01) and highly significant (P<0.001) differences.

IL-17-CD4 T cells specific for the S protein were only elevated in H-CoV and O-CoV groups, suggesting that infection was an inducer of Th17 responses (Fig. 4, G and H). O patients showed a non-significant trend towards increased IL-17 CD4 T cells compared to H donors (Fig. 4I). Equivalent results were obtained for the M protein (Fig. 4, J-L). Overall, these results indicated a stronger inflammatory response in O patients compared to H donors that could be associated to disease severity or cancer (fig. S2B). As expected from our previous results, inflammatory CD4 T cell subsets decayed 6 months after vaccination (Fig. 4, M and N).

Inflammatory S and M-specific CD8 T cell subsets were quantified (fig. S9B). Infection but not vaccination was the strongest inducer of S-specific IFNγ-CD8 T cells in H and O patients (Fig. 5, A-C). Similar results were obtained for M-specific T cells (Fig. 5, D and E), without differences between H donors and O patients (Fig. 5F). There were however marked differences for for IL-17-CD8 T cells, which were increased in subjects with previous COVID-19 following vaccination (Fig. 5, G and H). Although there was high variability, we observed a tendency towards increased IL-17-CD8 T cells in O patients compared to H donors (Fig. 5I). Similar results were obtained for M-specific CD8 T cells (Fig. 5, J and K), which were comparable between H donors and O patients (Fig. 5L). Although responses were in general low, inflammatory S- and M-specific CD8 T cells decayed 6 months after vaccination (Fig. 5, M and N).

**Fig. 5.**
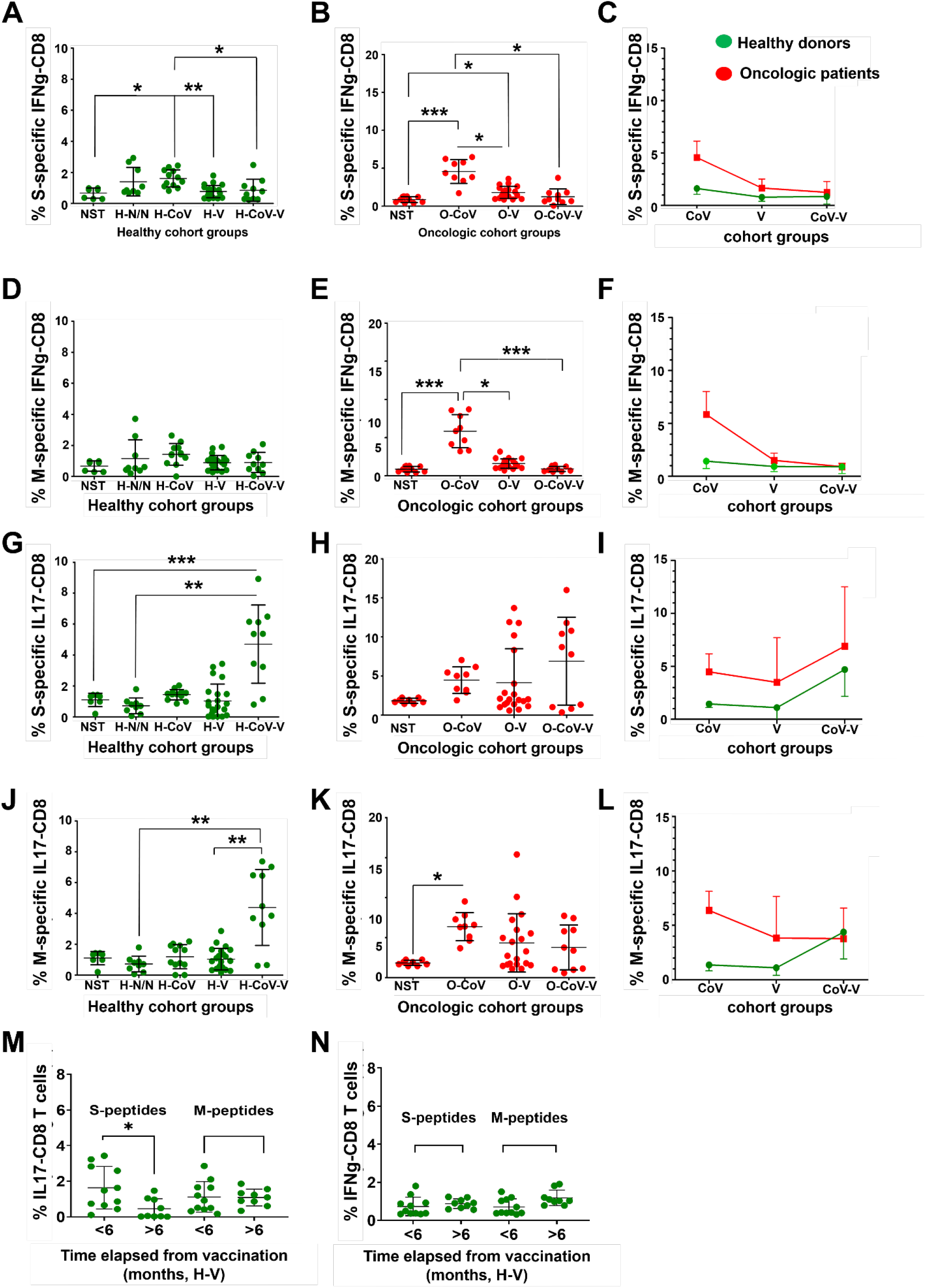
IFNγ and IL-17 expression in CD8 T cells specific for S and M proteins. **(A, B, C)** Percentage of IFNγ-CD8 T cells specific for the S protein in H and O groups. (D, E, F) Percentage of IFNγ-CD8 T cells specific for the M protein in H and O groups. (G, H, I) Percentage of IL-17-CD8 T cells specific for the S protein in H and O groups. (J, K, L) Percentage of IL-17-CD8 T cells specific for the M protein in H and O groups. (A-L) Statistical significance was evaluated with Kruskal-Wallis followed by Dunn’s pair-wise comparisons. (M, N) Percentage of CD8 T cells expressing IL-17 and IFNγ expression in H-V donors that completed the vaccine regime before sample collection in the indicated timelines, for both S and M proteins. Significance was tested with the U of Mann-Whitney test. NST, technical control of non-stimulated PMBCs. *, **, *** indicate significant (P<0.05), very significant (P<0.01) and highly significant (P<0.001) differences.

### Profiling of systemic myeloid cell subsets and B cells

The percentages of monocytes, granulocytes and neutrophils were quantified in peripheral blood and no differences were found in H donors (Fig. 6, A-C). However, there was a significant elevation of circulating granulocytes in O-CoV and O-CoV-V compared to O-V patients, suggesting that COVID-19 enhanced the expansion of systemic granulocytes (Fig. 6B). In contrast, O-V donors had significantly expanded the percentage of circulating monocytes, suggesting that vaccination targeted the monocytic lineage instead of the granulocytic lineage (Fig. 6, B-F). Hence, COVID-19 could perturb systemic immunity in cancer patients towards responses mediated by granulocytes rather than monocytes.

**Fig. 6.**
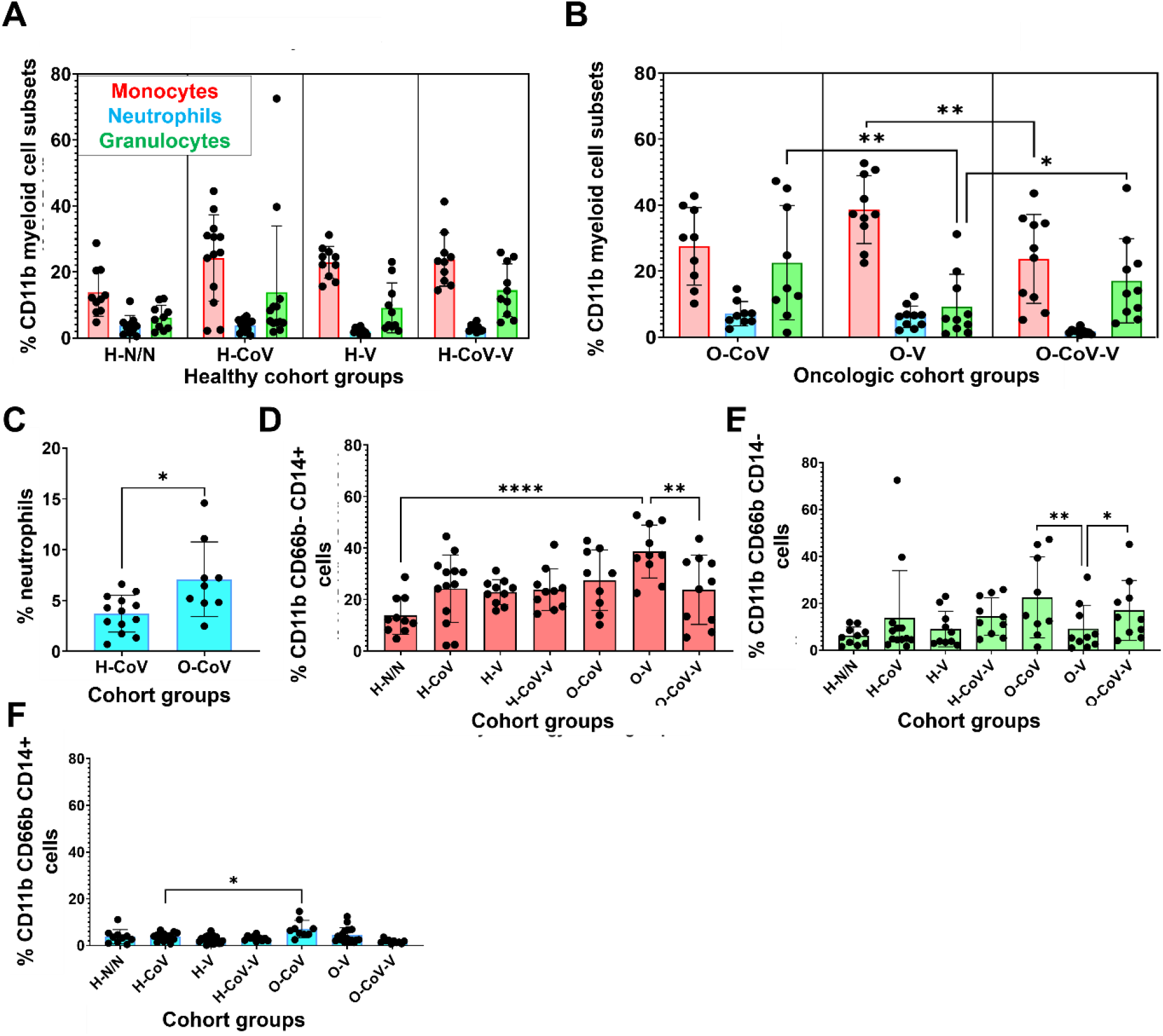
Systemic compositions of myeloid cell subsets. **(A, B)** Percentages of monocytes (CD11b+ CD66b-CD14+; red), neutrophils (CD11b+ CD66b+ CD14+; blue) and total granulocytes (CD11b+ CD66b+ CD14-, green), within CD11b+ cells. (C) Percentage of neutrophils in H-CoV and O-CoV. (D, E, F) Percentages of monocytes (CD11b+ CD66b-CD14+), neutrophils (CD11b+ CD66b+ CD14+) and granulocytes (CD11b+ CD66b+ CD14-) in the indicated groups. Statistical significance was evaluated by Kruskal-Wallis test followed by Dunn’s pair-wise comparisons. *, **, and *** indicate significant (P<0.05), very significant (P<0.01) and highly significant (P<0.001) differences.

Baseline percentages of circulating CD19+ CD14-B cells were also quantified, without finding significant differences (fig. S10, A-C). Nevertheless, there was a tendency in O patients to have decreased percentages of circulating B cells (fig. S10D).

## Discussion

Oncologic patients usually have a compromised immunity from cancer progression and treatments (10, 43) which may impact on responses to COVID-19 and vaccination. In this study we included S-, M- and N-specific T cell profiling and myeloid cell signatures. Most of donors had been vaccinated with mRNA BNT162b2. This vaccine is a potent inducer of S-specific antibodies (44-46) and we found that antibody responses were not impaired in cancer patients in agreement with others (47). In this study, we confirmed that antibody titres decreased over time which would limit serological protection to 6 months (48). Importantly, T cell responses after vaccination also decayed after 6 months. Indeed, vaccination did not preferentially expand memory T cell subsets, unless the subjects had previous COVID-19. Hence resolution of COVID-19 disease followed by vaccination may confer longer protection for both healthy donors and oncologic patients.

Vaccination mainly induced CD4 T cells, in contrast to SARS CoV-2 infection, which could explain its potency of raising antibody responses possibly through activated CD4 T helper cells (49). T cell responses towards M and S proteins were found previously (50, 51), but we extended this observation to O patients. In general, M protein was found to be a potent target for CD8 T cell responses even when compared to the S protein. Moreover, there was an expansion of CD4 T cells cross-reactive towards SARS CoV-2 M peptides. This is interesting, as protective cross-reactivity from COVID19 has already been observed by infection with common cold coronaviruses (51, 52). These results highlight the importance of including M-derived epitopes for developing novel vaccines.

O patients showed differences in T cell immunity compared to healthy donors. Their T cell repertoire was skewed towards differentiated phenotypes expressing IFNγ as shown before (23, 53), but also IL-17 as assessed here in H donors and O patients (54). Vaccination induced IFNγ and elevated IL-17 in CD4 T cells, a marker of Th17 responses (23, 53). Indeed, SARS-CoV-2 infection also induced a Th17 signature, which could be important for disease severity.

Finally, the profiles of circulating myeloid subsets was in agreement with oncologic patients having more inflammatory profile, as expected in cancer patients (43). This could be detrimental for vaccine efficacy. Elevated neutrophil counts are frequent in O patients (55, 56), which were even higher in subjects with previous COVID-19. A relationship between COVID-19 severity and higher monocyte and granulocyte content was found in early studies (57, 58).

Our study is not a controlled clinical trial but a retrospective analysis of immunological parameters with a limited number of patients, which could be a limitation. Nevertheless, we selected the sample size to ensure a minimum power of 0.8. Indeed, although the oncologic diseases were not homogeneous and included different solid tumors, the results were rather homogeneous which complemented the findings of other studies. Cancer patients showed proficient antibody, T cell and myeloid responses to infection and vaccination. Previous SARS CoV-2 infection had potent adjuvant effects for subsequent vaccination. These results suggest that vaccines such as those based on coronavirus replicons as priming (59) combined with conventional vaccines could be more efficacious than current vaccines. However, cancer patients showed baseline inflammation, which could be exacerbated upon infection followed by vaccination. Therefore, further studies on the consequences of vaccination and infection in cancer patients are merited.

## Materials and Methods

### Study cohort and design

This study was conducted according to the principles of the Declaration of Helsinki. The study was approved by the Clinical Research Ethics Committees of Hospital Universitario de Navarra and informed consents were obtained for all subjects. Study cohort and design are schematically depicted in supplementary figure S1. Peripheral blood samples from 57 healthy donors (H) and from 40 oncology patients (O) were obtained in the Oncology Unit of the University Hospital of Navarra (HUN), between April and December 2021. Samples corresponded to six study groups, including H donors and O patients with previous SARS CoV-2 infection (H-CoV, n=15; O-CoV, n=10), vaccinated without previous infection (H-V, n=22; O-V, n=20) and vaccinated after having an infection (H-V-CoV, n=10; O-V-CoV, n=10). SARS CoV-2 infection was confirmed by a positive PCR test. A group of H donors without previous infection nor vaccination was included as a control (H-N/N, n=10).

The total sample size of the study was established a priori to achieve a minimum power of 0.8 considering a large effect size (f=0.4) using Gpower 3.1 (40). We confirmed that sex did not have significant interaction with vaccination in either healthy donors or oncologic patients in any of the tested variables. General clinical characteristics and SARS CoV-2-related parameters of the study cohort are summarized in Table 1 and Table 2, respectively. Infected patients were classified for COVID-19 severity according to the Treatment Guidelines of the NIH (https://www.covid19treatmentguidelines.nih.gov/overview/clinical-spectrum/):

0 = Asymptomatic infection: Positive for SARS-CoV-2 without symptoms.

1 = Mild illness: Any of the symptoms of COVID-19 without shortness of breath, dyspnoea, or abnormal chest imaging.

2 = Moderate illness: Evidence of lower respiratory disease during clinical assessment or imaging with oxygen saturation (SpO2) ≥94% on room air at sea level.

3 = Severe illness: Individuals with SpO2 <94%, a ratio of arterial partial pressure of oxygen to fraction of inspired oxygen (PaO2/FiO2) <300 mm Hg, a respiratory rate >30 breaths/min, or lung infiltrates >50%.

4 = Critical illness: Respiratory failure, septic shock, and/or multiple organ dysfunction.

### Sample processing, PBMCs reactivation and flow cytometry

Blood collection, PBMC, myeloid cells and T cell purification, activation and flow cytometry were carried out as previously described (10). The following fluorochrome-conjugated antibodies were used: CD14-Violet Fluor 450 (Ref 75-0149-T100, TONBO), CD11b-PerCP-Cy5-5 (Ref 65-0112-U1, TONBO), CD62L-APC (Ref 130-113-617, Miltenyi), CD66b-APC-Cy7 (Ref 130-120-146, Miltenyi), CD54-FITC (Ref 130-104-214, Miltenyi), CD19-PE (Ref 130-113-731, Miltenyi) , CD3-APC (Ref 130-113-135, Miltenyi), CD8-APC-Cy7 (Ref 130-110-681, Miltenyi), CD4-FITC (Ref 130-114-531, Miltenyi), CD27-PE (Ref 50-0279-T100, TONBO), CD28-PE-Cy7 (Ref 130-126-316, Miltenyi). CD8-PE-Cy7 (Ref 130-110-680, Miltenyi), CD4-APC-Cy7 (Ref 25-0049-T100, TONBO), CD154-PerCP-Cy5-5 (Ref 130-122-800, Miltenyi), CD137-PE (Ref 130-110-763, Miltenyi), IFNγ-FITC (Ref 130-113-497, Miltenyi), IL-17-Vio770 (Ref 130-118-249, Miltenyi), CD45RA-FITC (Ref 35-0458-T025, TONBO), CD62L-APC (Ref 130-113-617, Miltenyi).

For T cell activation half a million PBMCs were plated per well in a 96-well plate, and reactivated with 0.8 ng/μl of the following SARS-CoV-2 PepTivators (Miltenyi) separately: PepTivator SARS-CoV-2 Prot_M, PepTivator SARS-CoV-2 Prot_N, PepTivator SARS-CoV-2 Prot_S, PepTivator SARS-CoV-2 Prot_S1, PepTivator SARS-CoV-2 Prot_S+. S protein PepTivators were mixed. Cells were incubated for 17-19 hours at 37°C, and then treated with 1 μl/ml of Brefeldin A (ThermoFisher Scientific). Cells were washed and stained for flow cytometry.

### SARS CoV-2 protein expression and purification

For ELISA and stimulation studies M, S and N proteins were produced using Bac-to-Bac baculovirus expression. Briefly, synthetic genes encoding S1 (1-303 amino acid), full length N and the cytoplasmic domain of the M protein (1-100 amino acid) were fused to histidine tags and cloned. Protein production and purification by Ni-NTA affinity and size exclusion chromatographies were performed following standard protocols (Bac-to-Bac Thermofisher).

### Non-sandwich ELISA

Donor sera were obtained from peripheral blood, centrifuged and frozen at -20ºC. For detection of S and N specific antibodies, a 96-well plate was coated with 5 μg/mL of the corresponding protein, followed by blocking with 1X PBS-2% BSA. 1:800, 1:250 and 1:80 sera dilutions were used for detection of anti-S antibodies, anti-N antibodies and anti-M antibodies, respectively. Anti-human IgGs HRP-labelled antibody (ThermoFisher) was used as secondary antibody. ELISAS were developed with 100 μL TMB substrate (Sigma) and read at 450 nm.

#### Statistical analyses

Statistical analyses were performed with GraphPad 8. Variables under study were tested for normality (Kruskal-Wallis test), homogeneity of variances (F test), and homogeneity (Spearman’s coefficient of variation). Antibody titres and percentages of cell types as quantified by flow cytometry were either not normally distributed or showed high variability. For multi-group comparisons of these variables, non-parametric Kruskal-Wallis tests were performed followed by pair-wise comparisons with Dunn’s test. For experiments involving only two independent groups, the non-parametric U of Mann Whitney was used. The percentages of T cell phenotypes were normally distributed, homogeneous and with comparable variances. In this case, one-way ANOVAs were carried out followed by a posterori pair-wise comparisons with Tukey’s test.

## Data Availability

All data produced in the present work are contained in the manuscript.

## Author contributions

Conceptualization: GK, DE, MA, RV

Methodology: ME, MD, AFL, PO, GK, DE, MA, RV

Investigation: ME, MG, PR, LC, LF, HA, AB, EB, SP

Visualization: ME, IL, MG, PR, LC, LF, HA, AB, EB, SP, GK, DE, MA

Supervision: GK, DE, MA, RV

Writing—original draft: ME, GK, DE

Writing—review & editing: ME, GK, DE, IB, MA

## Data and materials availability

All data are available in the main text or the supplementary materials.

---

**Fig. S1.**
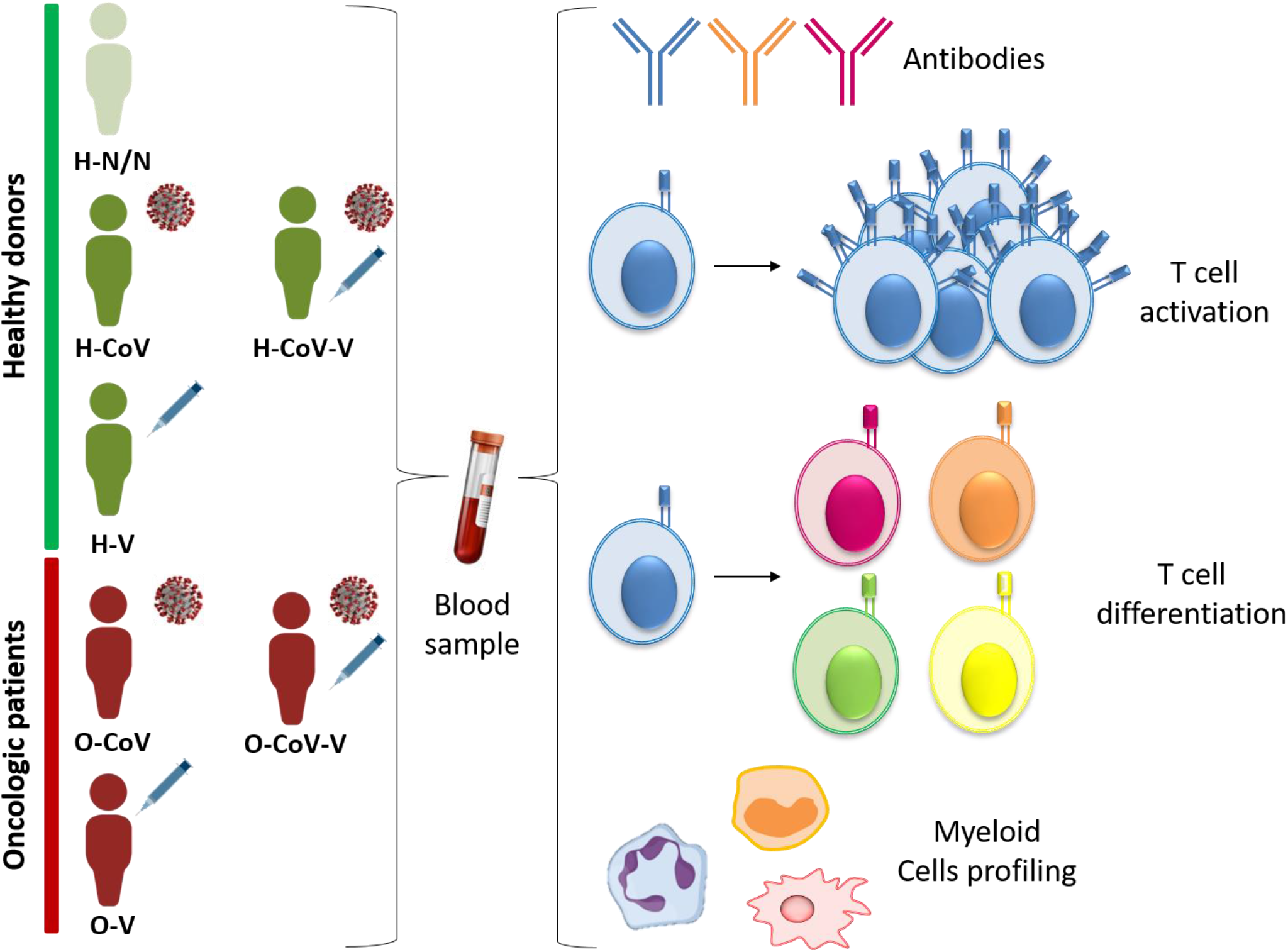
Schematic representation of the study cohort and design. Samples from H donors and O patients naturally infected by SARS CoV-2 and vaccinated with/without previous SARS CoV-2 infection were collected. A group of non-infected non-vaccinated H donors was included for comparison. Blood samples were obtained and analysed for antibody levels, T cell activation and differentiation, and myeloid cell characterisation.

**Supplementary figure S2.**
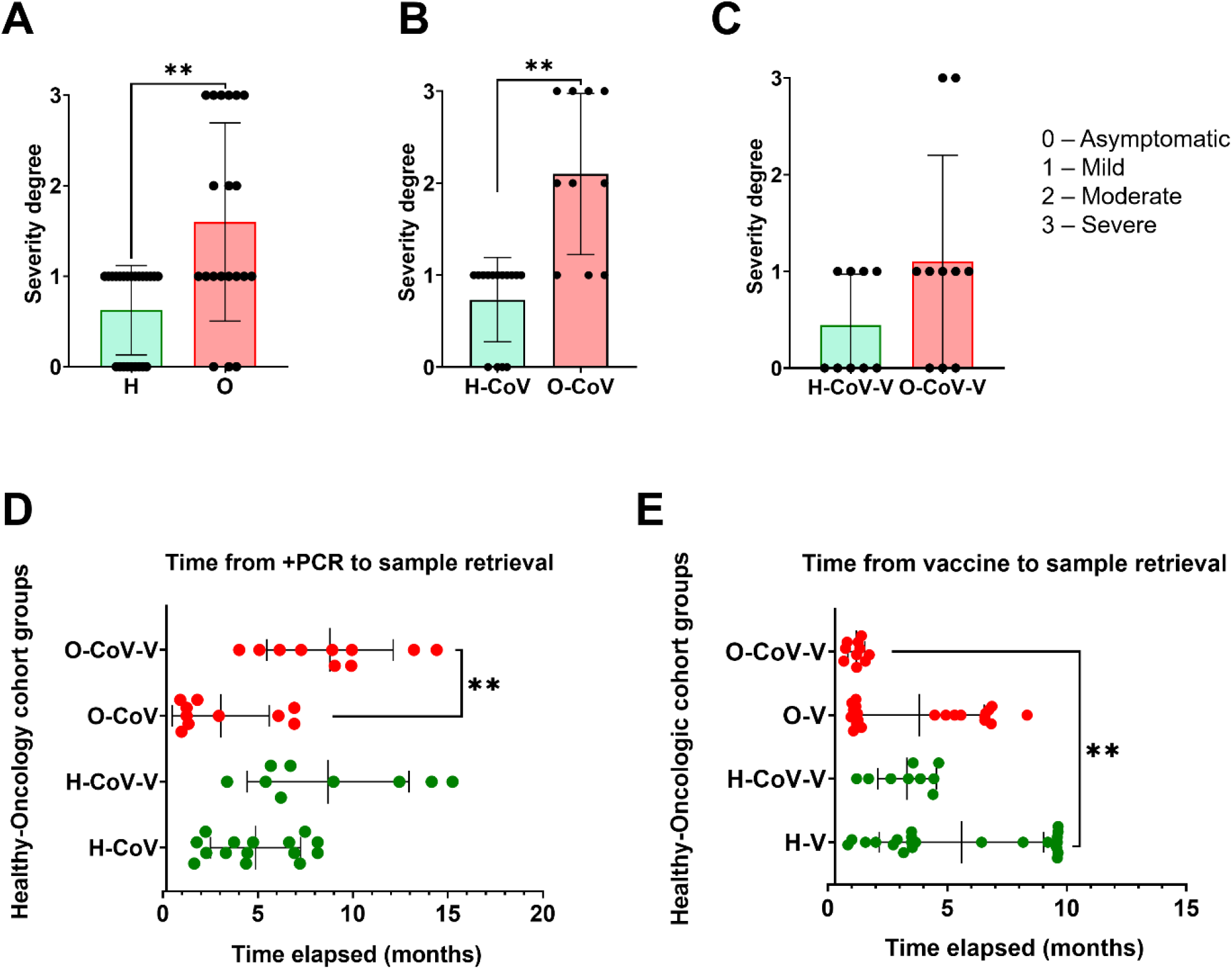
COVID-19 severity degree and time elapsed from SARS CoV-2 infection and/or vaccination to sample. **(A, B, C)** COVID-19 severity degree according to the NIH guidelines in all H and O individuals, in H-CoV and O-CoV and in H-CoV-V and O-CoV-V. The U of Mann-Whitney test was used to evaluate significance. **(D, E)** Time elapsed from SARS CoV-2 diagnosis to sample collection and from vaccination to sample collection in the indicated groups of H donors and O patients. Kruskal-Wallis test was used for multiple comparisons followed by Dunn’s test for pair-wise comparison. *, **, indicate significant (P<0.05) and very significant (P<0.01) differences.

**Fig. S3.**
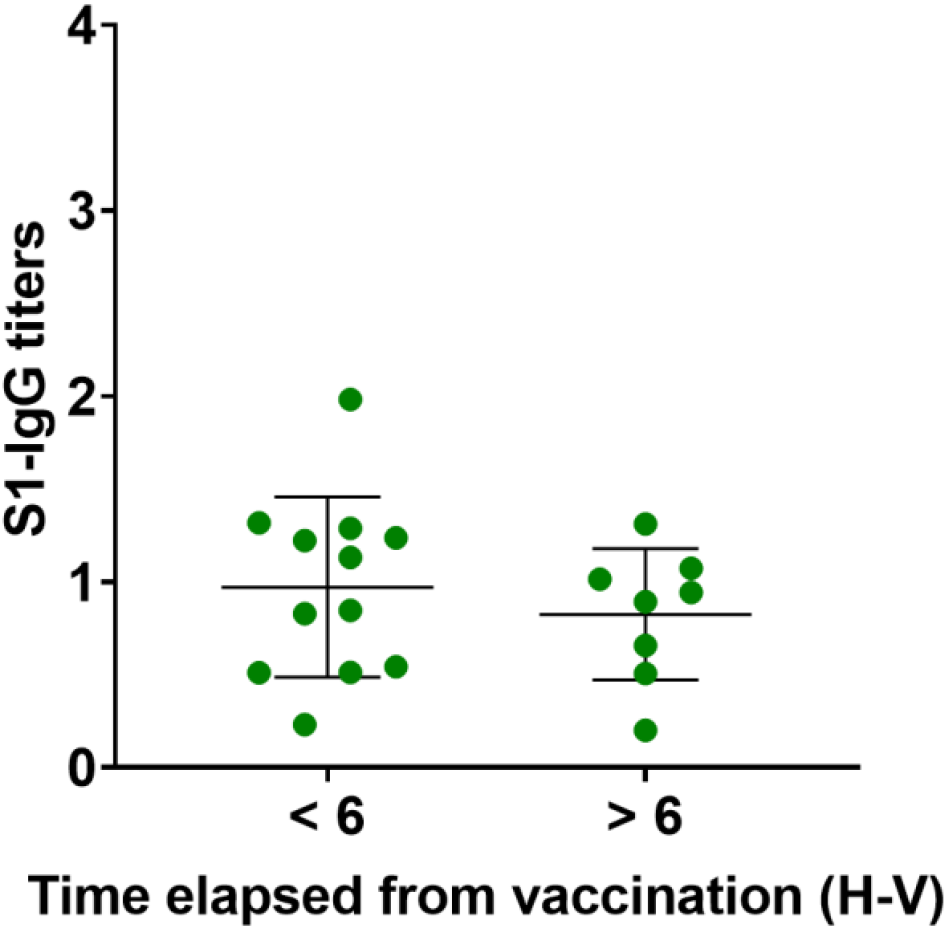
Dynamics of S-specific IgG titres. S specific IgG antibody titres in V-H individuals from samples collected less than 6 months and more than 6 months after vaccination. The U of Mann-Whitney test was used for statistical significance.

**Fig. S4.**
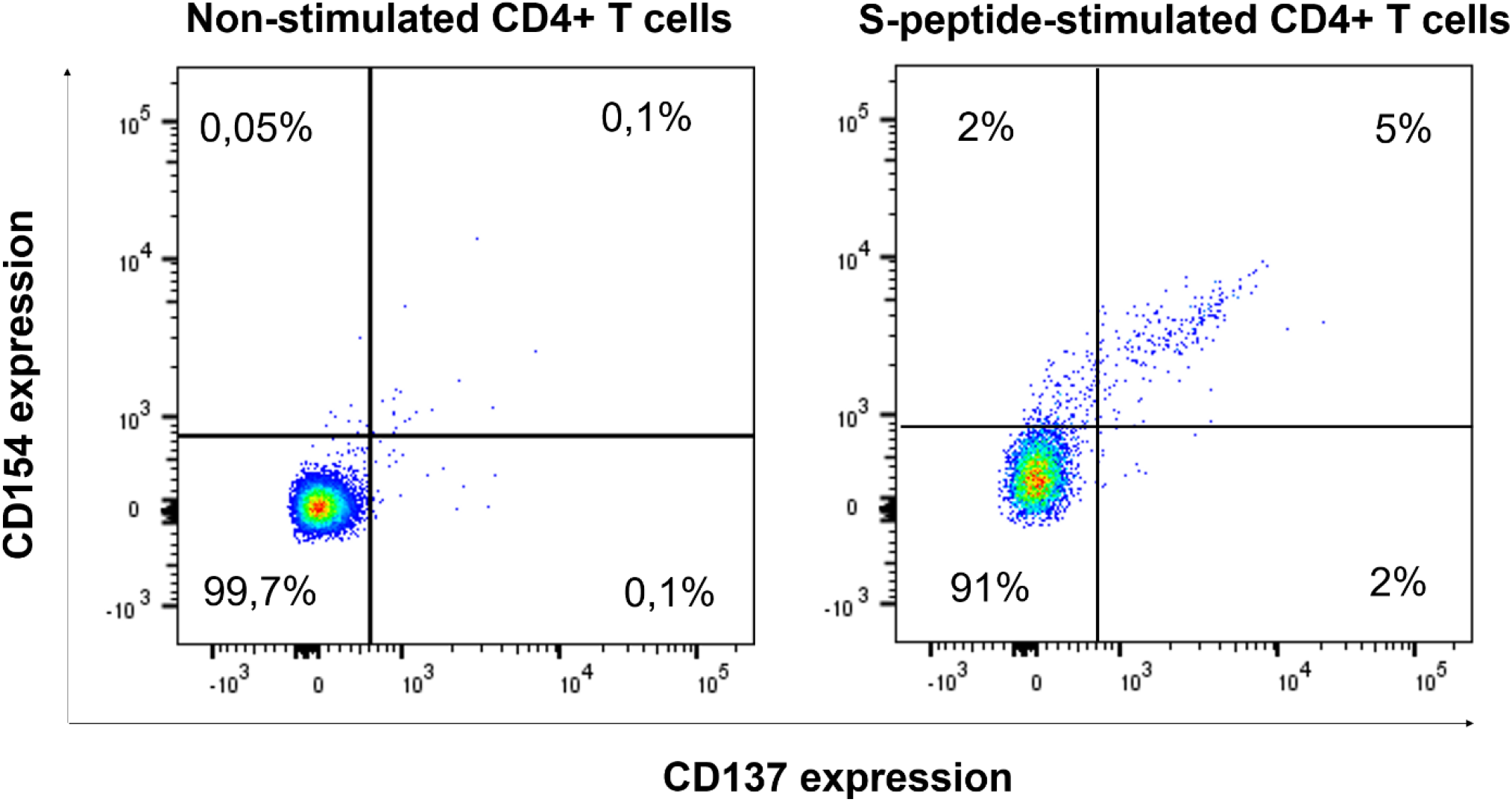
Flow cytometry and gating strategy for quantification of activated CD4 T cells. Representative flow cytometry density plots of CD154 and CD137 co-expression profiles in CD4 T cells from donors before and after stimulation with S-peptides, as indicated.

**Fig. S5.**
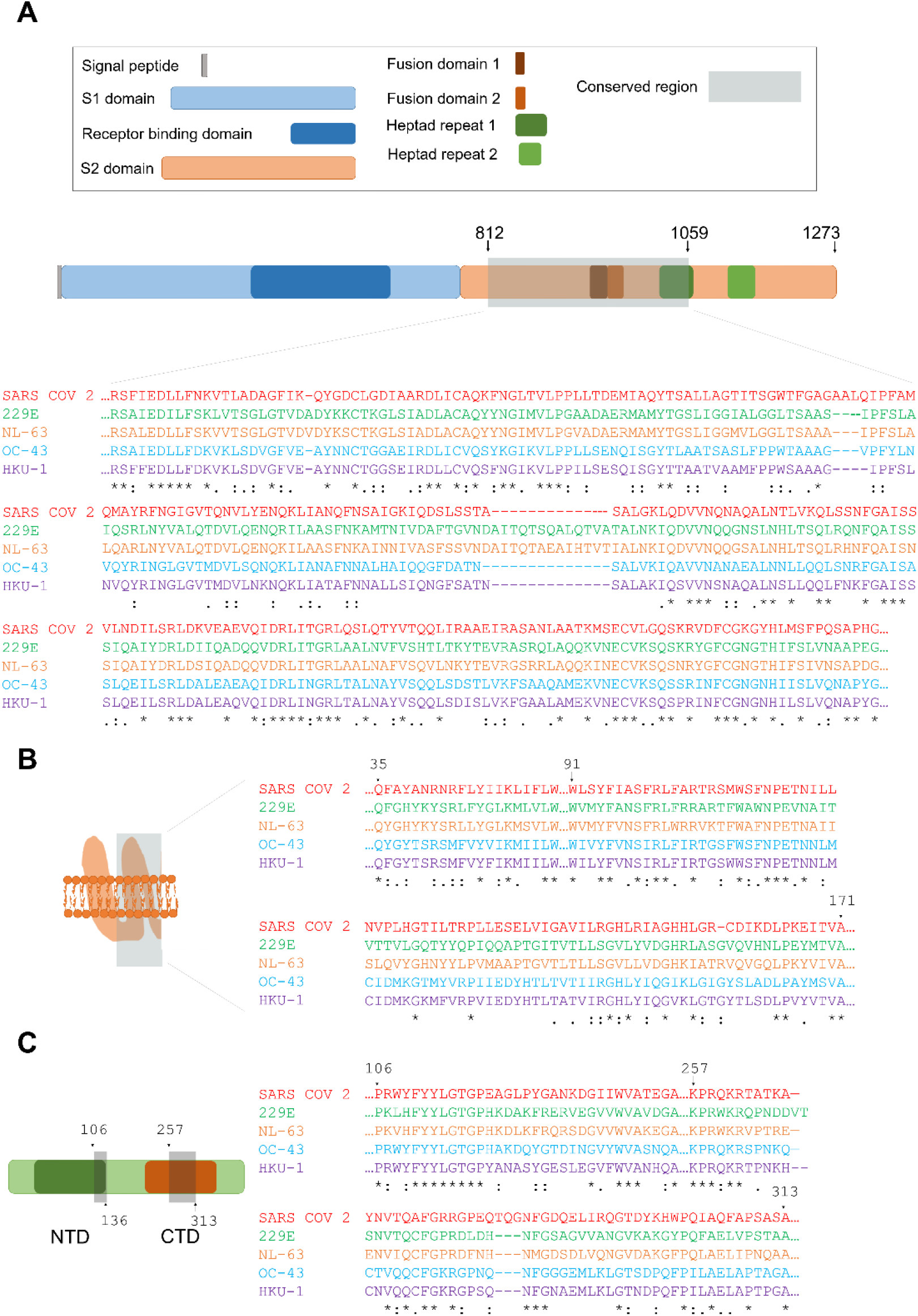
Sequence alignment. **(A, B, C)** Conserved region of the spike protein, membrane protein and nucleocapsid protein of SARS-CoV-2 (in red) with the indicated human coronavirus counterparts: 229E (in green), NL-63 (in orange), OC-43 (in blue) and HKU-1 (in purple). Conserved regions are highlighted in grey.

**Supplementary figure S6.**
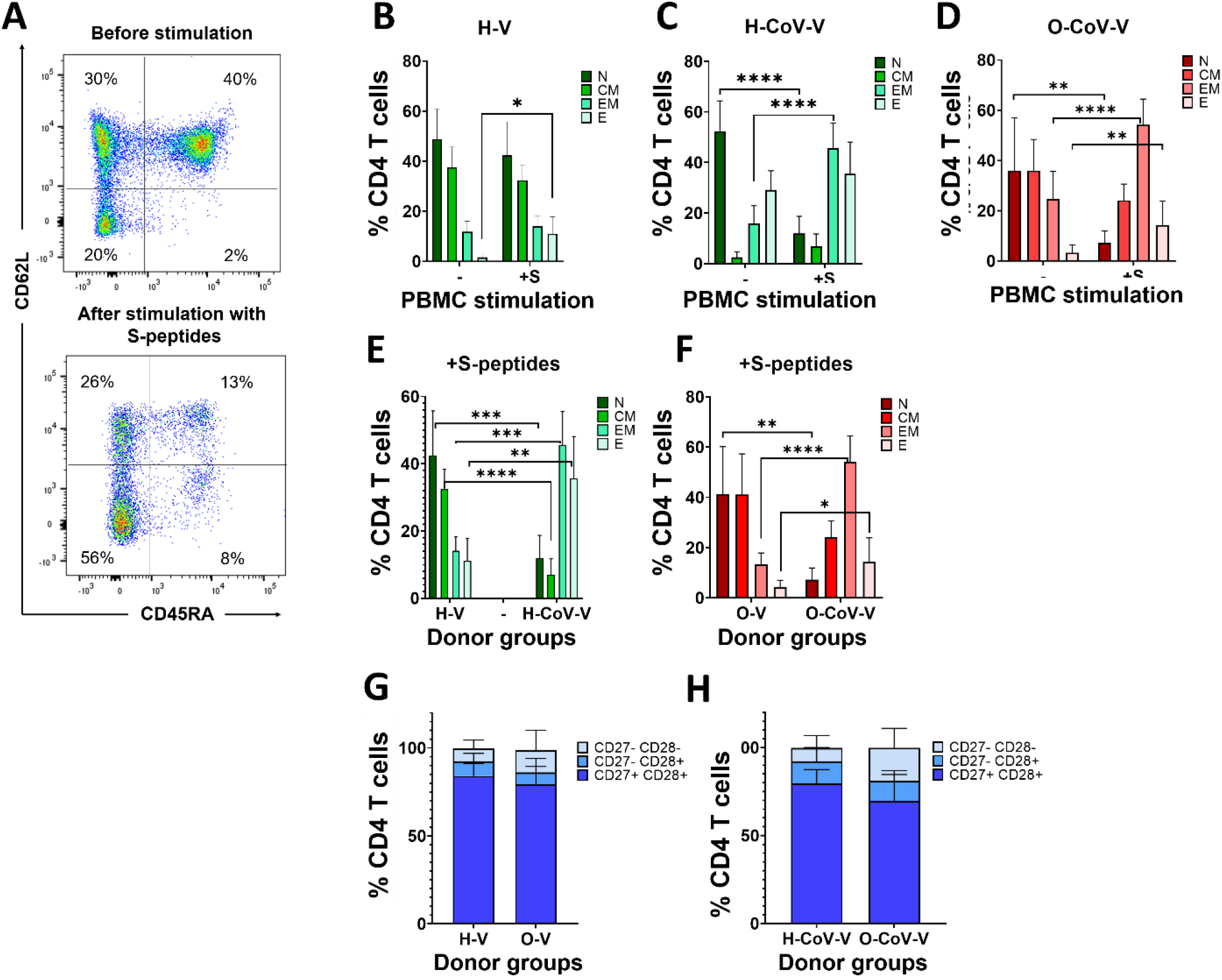
CD4 T cell phenotypes after stimulation with S peptides. **(A)** Representative flow cytometry density plots with CD62L-CD45RA co-expression profiles in CD4 T cells before and after the stimulation with S-peptides. Quadrants were established with unstained controls. Percentages of the corresponding populations are shown within the quadrants. **(B, C)** CD4 T cell phenotypic changes in H-V and H-CoV-V donors. (-) and (+S), non-stimulated and S-peptide stimulation. N, CM, EM and E, indicate naïve-stem cell (CD62L+ CD45RA+), central memory (CD62L+ CD45RA-), effector memory (CD62L-CD45RA-) and effector (CD62L-CD45RA+) phenotypes. **(D)** Phenotypic changes in CD4 T cells within O-CoV-V before and after stimulation with S-peptides. **(E**,**F)** Effects of previous CoV infection in vaccinated H donors and O patients over T cell phenotypes after stimulation with S-peptides. (B-F) Relevant statistical comparisons are indicated by ANOVA followed by pair-wise comparisons with Tukey’s test. **(G, H)** Relative percentages of CD4 T cell differentiation phenotypes in H-V and O-V and in H-CoV-V and O-CoV-V CD27+ CD28+, CD27-CD28+ and CD27+ CD28+ indicate poorly differentiated, intermediate differentiated and highly differentiated T cell phenotypes. U of Mann-Whitney was used to test for significance.*, **, *** indicate significant (P<0.05), very significant (P<0.01) and highly significant (P<0.001) differences.

**Supplementary figure S7.**
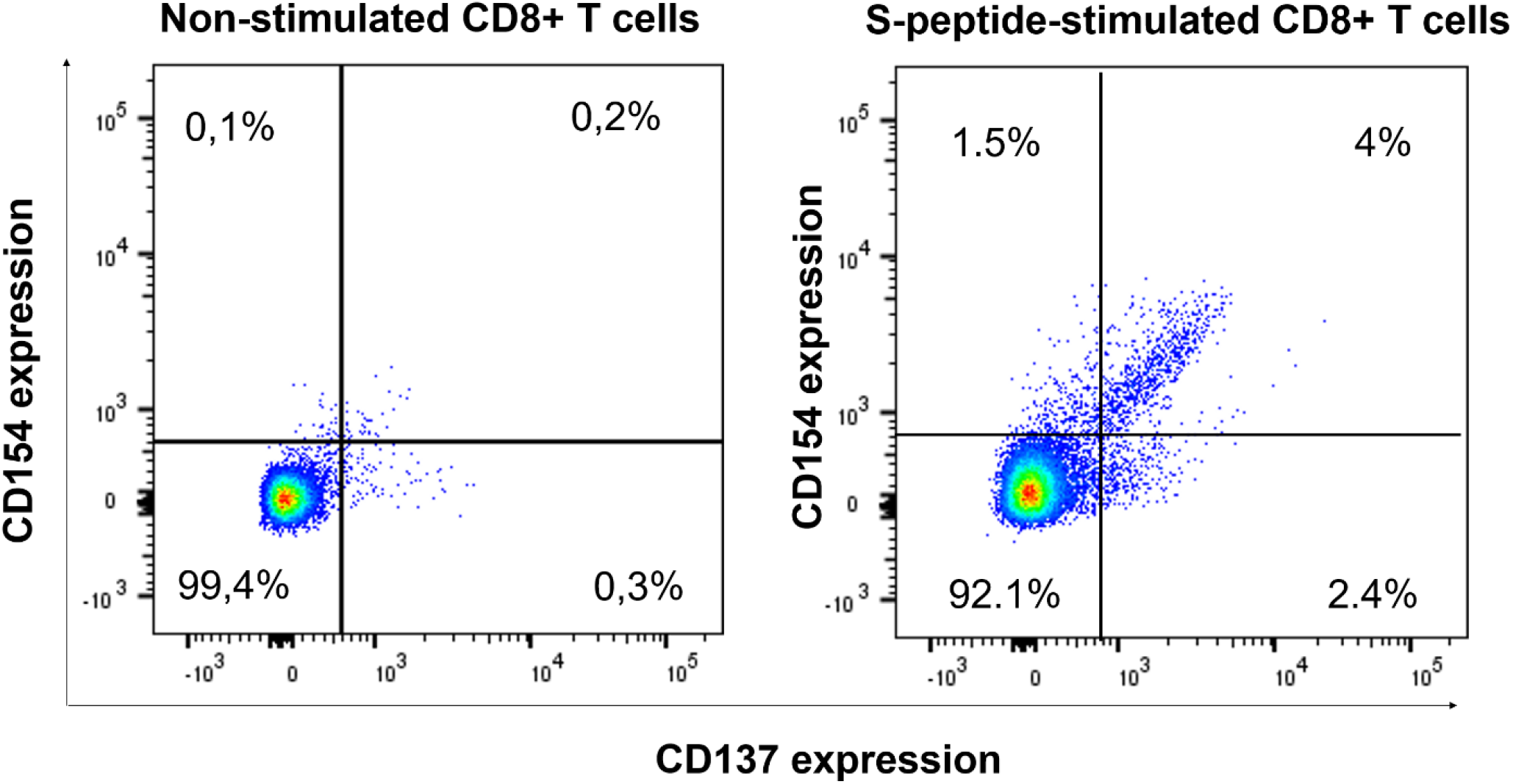
Flow cytometry and gating strategy for quantification of activated CD8 T cells. Representative flow cytometry density plots of CD154 and CD137 co-expression profiles in CD8 T cells from donors before and after stimulation with S-peptides, as indicated.

**Fig. S8.**
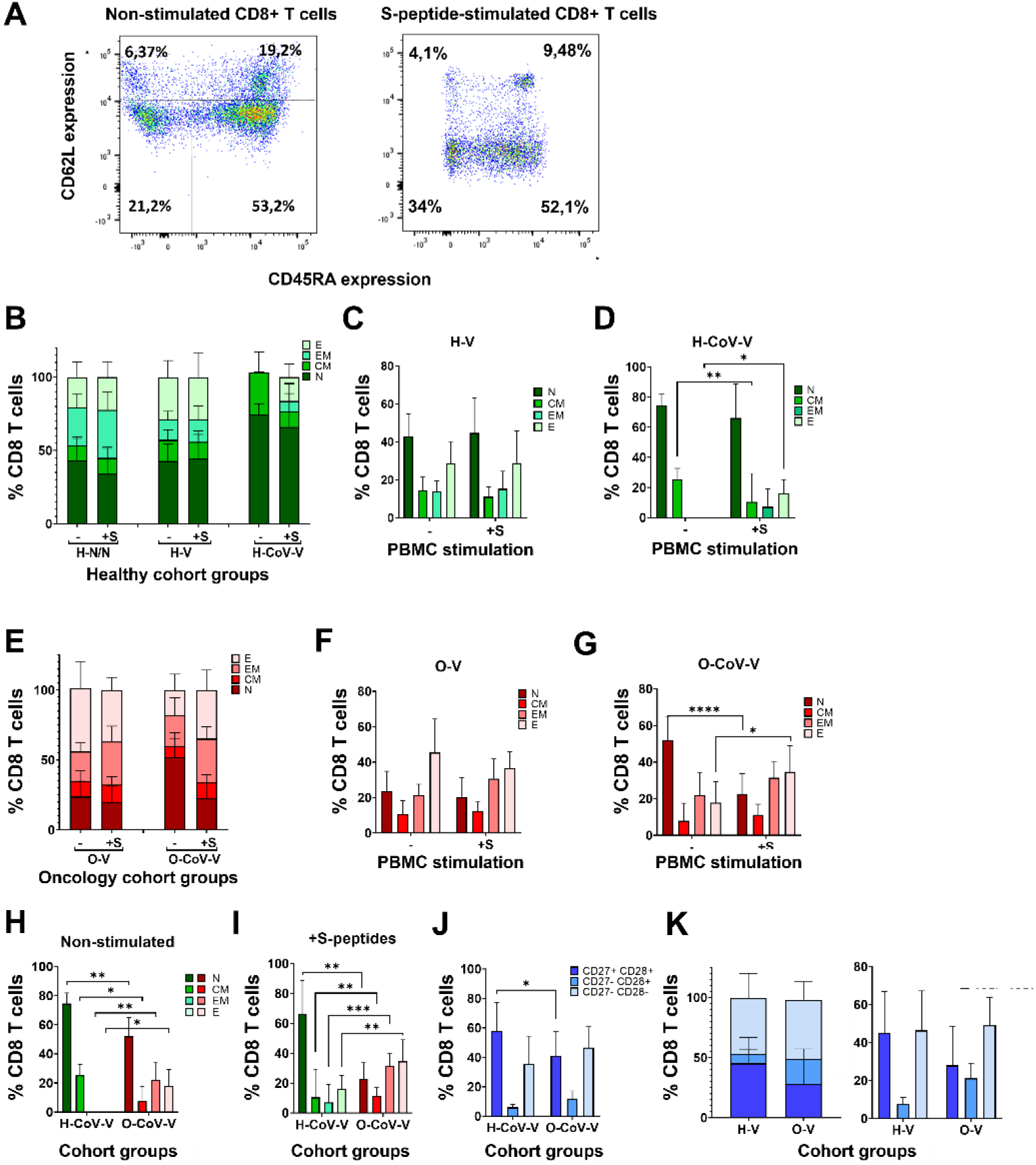
Differentiation phenotypes in CD8 T cells. **(A)** Representative flow cytometry density plots with CD62L-CD45RA co-expression profiles in CD8 T cells before and after the stimulation with S-peptides. Quadrants were established with unstained controls. Percentages of the corresponding populations are shown within the quadrants. **(B, C, D)** Relative percentages of CD8 T cell differentiation phenotypes from the indicated H donors and O patient cohorts. Means and error bars (standard deviations) are shown. N, CM, EM and E, indicate naïve-stem cell (CD62L+ CD45RA+), central memory (CD62L+ CD45RA-), effector memory (CD62L-CD45RA-) and effector (CD62L-CD45RA+) phenotypes. **(E, F, G)** Relative percentages of CD8 T cell differentiation phenotypes from the indicated H and O cohort groups. **(H**,**I)** Relative percentages of CD8 T cell differentiation phenotypes in H-CoV-V and O-CoV-V groups before and after stimulation with S-peptides. **(J, K)** Relative percentages of CD8 T cell differentiation phenotypes in V-H-CoV, V-O-CoV, H-V and O-V groups as indicated in the graphs. CD27+ CD28+, CD27-CD28+ and CD27+ CD28+ indicate poorly differentiated, intermediate differentiated and highly differentiated T cell phenotypes. (B-K) Statistical significance was tested by ANOVA followed by Tukey’s pair-wise comparisons. *, ** and *** indicate, significant (P<0.05), very significant (P<0.01) and highly significant (P<0.001) differences.

**Fig. S9.**
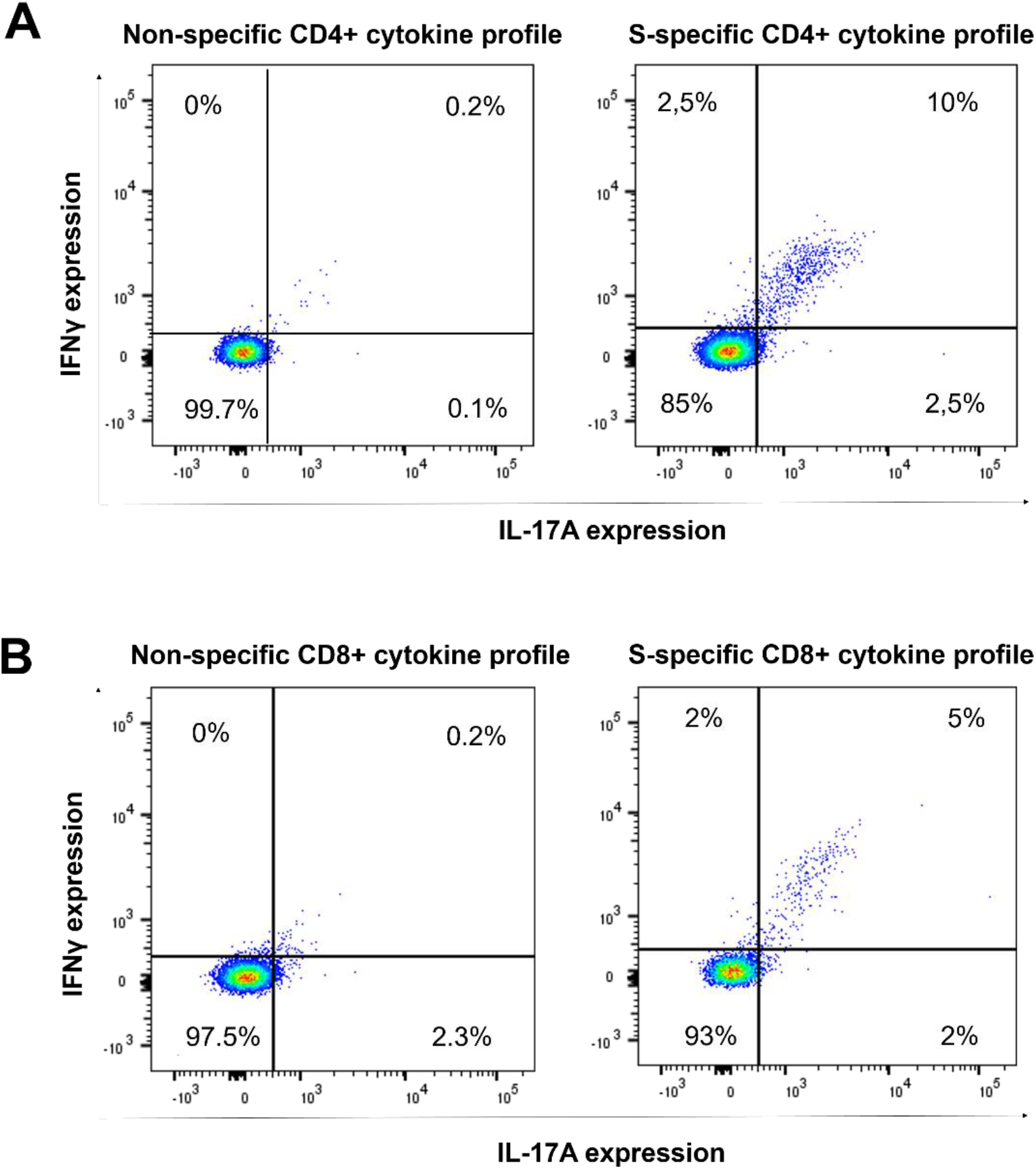
Flow cytometry and gating strategy for quantification of cytokine expression within activated T cells. **(A)** Representative flow cytometry density plots with the expression of INFγ and IL-17 before and after stimulation with S peptides in CD4 T cells. **(B)** in CD8 T cells.

**Fig. S10.**
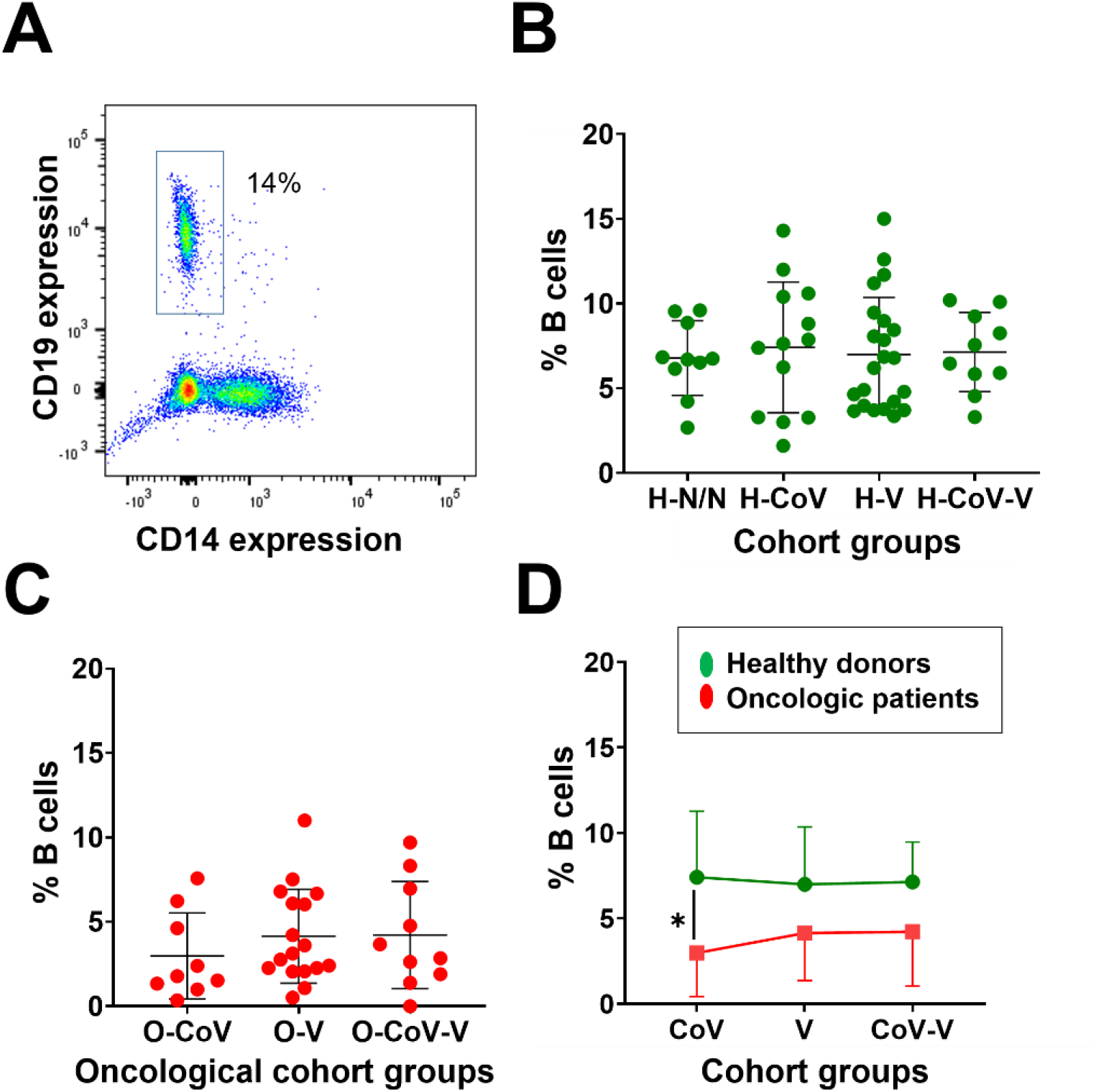
Quantification of circulating B cell in peripheral blood from H donors and O donors. **(A)** Representative flow cytometry density plot of CD19 and CD14 expression in PBMCs from donors. **(B, C)** Percentage of circulating B-cells in the indicated groups of H donors and O patients. **(D)** Percentage of circulating B-cells in the indicated groups of H (in green) and O (in red) subjects. Statistical significance was tested with Krustal-Wallis followed by Dunn’s pair-wise comparisons. *, **, and *** indicate significant (P<0.05), very significant (P<0.01) and highly significant (P<0.001) differences.

